# Prognostic peripheral blood biomarkers at ICU admission predict COVID-19 clinical outcomes

**DOI:** 10.1101/2022.01.31.22270208

**Authors:** Melina Messing, Mypinder S. Sekhon, Michael R. Hughes, Sophie Stukas, Ryan L. Hoiland, Jennifer Cooper, Nyra Ahmed, Mark Hamer, Yicong Li, Samuel B. Shin, Lin Wei Tung, Cheryl Wellington, Don D. Sin, Kevin B. Leslie, Kelly M. McNagny

**Affiliations:** The Biomedical Research Centre, School of Biomedical Engineering, University of British Columbia; Vancouver, BC, Canada; Department of Medicine (Division of Respirology), University of British Columbia; Vancouver, BC, Canada; Department of Pathology and Laboratory Medicine, University of British Columbia; Vancouver, BC, Canada; Department of Anesthesiology, Pharmacology and Therapeutics, University of British Columbia; Vancouver, BC, Canada; Centre for Heart Lung Innovation (HLI), University of British Columbia; St Paul’s Hospital, Vancouver, BC, Canada

**Keywords:** COVID-19, ICU admission, immune biomarkers

## Abstract

The COVID-19 pandemic continues to challenge the capacities of hospital ICUs which currently lack the ability to identify prospectively those patients who may require extended management. In this study of 90 ICU COVID-19 patients, we evaluated serum levels of four cytokines (IL-1β, IL-6, IL-10 and TNFα) as well as standard clinical and laboratory measurements. On 42 of these patients (binned into Initial and Replication Cohorts), we further performed CyTOF-based deep immunophenotyping of peripheral blood mononuclear cells with a panel of 38 antibodies. All measurements and patient samples were taken at time of ICU admission and retrospectively linked to patient clinical outcomes through statistical approaches. These analyses resulted in the definition of a new measure of patient clinical outcome: patients who will recover after short ICU stays (< 6 days) and those who will subsequently die or recover after long ICU stays (≥ 6 days). Based on these clinical outcome categories, we identified blood prognostic biomarkers that, at time of ICU admission, prospectively distinguish, with 91% sensitivity and 91% specificity (positive likelihood ratio 10.1), patients in the two clinical outcome groups. This is achieved through a tiered evaluation of serum IL-10 and targeted immunophenotyping of monocyte subsets, specifically, CD11c^low^ classical monocytes. Immunophenotyping revealed clear predictors of clinical outcome in COVID-19 providing a highly sensitive and specific prognostic test that could prove useful in guiding clinical resource allocation.

**Graphical Abstract:** 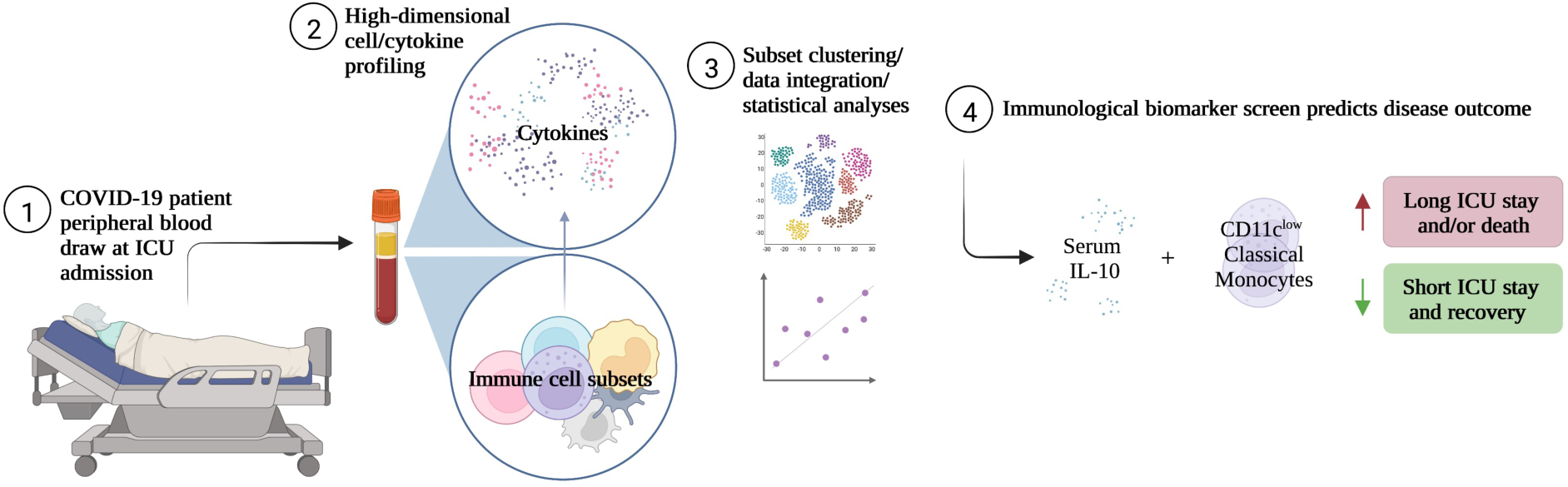

## Introduction

COVID-19 continues to overwhelm effective health care delivery in most parts of the world due to challenges in achieving sufficiently high vaccination rates, vaccine hesitancy and the emergence of more transmissible viral variants for which current vaccines offer more modest protection. Thus, waves of rapid outbreaks continue to threaten ICU capacities (1, 2).

Individual patient responses to infection by the SARS-CoV-2 virus can vary dramatically, ranging from asymptomatic or mild flu-like symptoms to much more severe symptoms including acute respiratory distress syndrome (ARDS) and death (3, 4). Individual patient outcomes are remarkably challenging to predict but severe disease has been broadly linked to advanced age, obesity, underlying comorbidities and secondary infections (5–10). Neither symptoms nor conventional clinical laboratory measurements (serum C-reactive protein (CRP), blood D-dimers etc.) have sufficient prognostic power and, thus, approved interventions for severe COVID-19 (including systemic corticosteroids and tocilizumab) are administered broadly to patients admitted to the ICU as clinicians lack the tools to identify accurately patients at risk of long-term complications and death (11–15). Immunologically, “severe” COVID-19 patients have been reported to exhibit lymphopenia, neutrophilia, accumulation of lung monocytes, emergency myelopoiesis and substantial changes in serum cytokine and chemokine profiles likely reflecting a cytokine storm as the result of a delayed, but exuberant, immune response to infection (16–23). The latter has been of particular interest for the development of prognostic tools but, while some markers have proven useful in measuring the severity of active COVID-19, to date they have lacked the necessary statistical power to prospectively predict the likelihood of incipient severe disease (24–31). For example, serum IL-6 (alone, or together with other inflammatory markers) has most consistently been linked to severe active disease and, by some groups, was shown to predict the need for subsequent mechanical ventilation as well as survival (32–36). In contrast, other studies have struggled to link tightly serum IL-6 (or TNFα, IFNγ or GM-CSF) to an elevated risk of severe disease and instead have proposed various combinations of serum levels of CCL5, IL-1RA and IL-10, EN-RAGE, TNFSF14 and oncostatin M as indicators of incipient severe disease and, in some cases, predictors of disease severity (37–39). While, individually, these studies show several biomarkers capable of triaging patients, the inconsistent and sometimes contradictory results highlight the need for biomarkers with robust statistical power to be clinically useful.

Studies focused on cellular changes have linked a decreased frequency of monocytes (and, more variably, alterations in the frequency of natural killer cells (NK), plasmacytoid dendritic cells (pDC), type-2 conventional dendritic cells (DC2), mucosal associated invariant T (MAIT) cells and other lineages) with active severe disease and poor outcomes (17, 19, 34, 40–44). While these global cellular profiling efforts provide important insights into the immune response to SARS-CoV-2 infection, they have yet to be translated into prognostic tools to assist with individualized care.

Here we focused on the development of an immunological biomarker screen that, at ICU admission for COVID-19, predicts length of ICU stay or death. Strikingly, we find that, at ICU admission, measurements of serum IL-10 and simple monocyte subset surface signatures, specifically, CD11c^low^ classical monocytes, can predict with 91% sensitivity and 91% specificity patients who will either die or have a longer stay in ICU. We offer these biomarkers as a model clinical laboratory test with future potential in gaining insights into variable responses to SARS-CoV-2 infection.

## Results

### COVID-19 patient group selection and optimization of high dimensional immune profiling

To identify potential prognostic markers of COVID-19, we collected, within the first 48 hours of ICU admission, serum samples from 90 ICU COVID-19 patients (the “Cytokine Cohort”) admitted during the “second wave” of COVID-19 (November, 2020–February, 2021) together with serial serum samples across different timepoints during the course of their ICU admission. PBMCs collected from 14 of these 90 ICU COVID-19 patients (the “Initial Cohort”) were analyzed by mass cytometry (CyTOF) with a training set of 35 monoclonal antibodies (Supplementary Table 1) designed to detect broad shifts in levels of normal PBMC lineages as well as their activation status and the possible mobilization of tissue resident innate immune cells and bone marrow progenitors into peripheral blood. Based on these data we developed a refined, second-generation, 38-antibody CyTOF panel (Supplementary Table 1), which was used on PBMCs collected from a further 28 of the 90 ICU COVID-19 patients (the “Replication Cohort”). Data from the Replication Cohort were used to validate observations from the Initial Cohort on early alterations in immune responses that could effectively differentiate patients likely to recover after a short ICU stay from those who would either die or have prolonged stays in ICU. The ICU admission sera from all patients in the Cytokine Cohort (which includes all patients in the Initial Cohort and the Replication Cohort) were analyzed for levels of four cytokines: IL-1β, IL-6, IL-10 and TNFα (Fig. 1A, B). PBMC, but not serum samples, were obtained for 8 healthy, age-matched controls for the CyTOF analyses portion of this study.

**Fig. 1.**
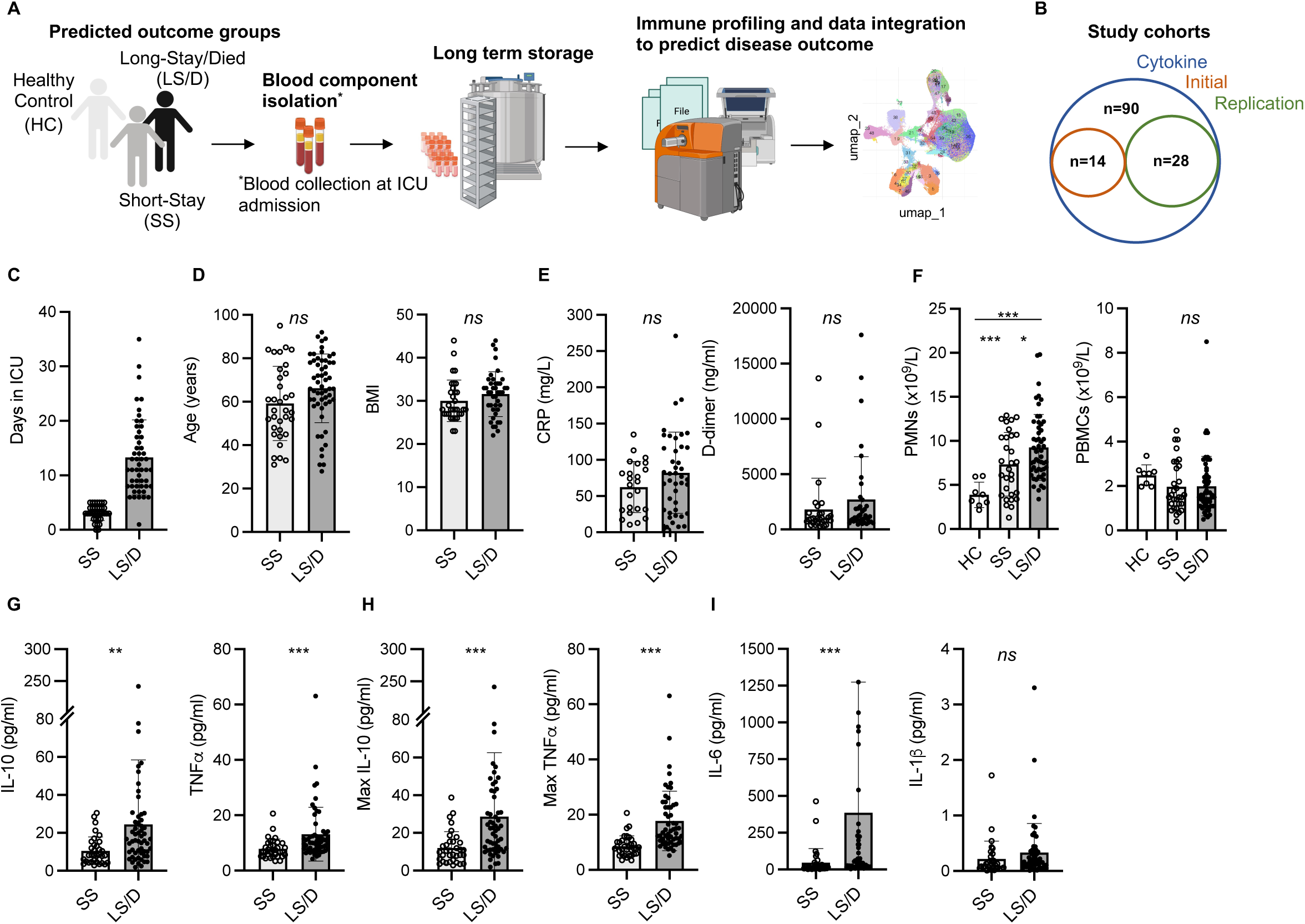
Patient cohort selection, characteristics and cytokine analyses. Experimental design overview: peripheral blood was collected from COVID-19 patients within 48h of ICU admission; immune cells and serum were isolated and stored followed by immune cell subset and cytokine analyses and clinical data integration (**A**). Patient cohorts overview; Initial Cohort SS patient n = 7, LS/D patient n = 7; Replication Cohort SS patient n = 12, LS/D patient n = 16 (**B**). Patient outcome groups based on length of stay in ICU (C). Patient age, body mass index (BMI), C-reactive protein (CRP) levels and D-dimer levels (**D-E**). Complete blood counts of patients and healthy controls (**F**). Serum cytokine levels of IL-10, TNFα, maximum IL-10, maximum TNFα, IL-6 and IL-1β (**G-I**). *, p < 0.05; **, p < 0.01; ***, p < 0.001; ns, p ≥ 0.05 by two-tailed, two-sample unequal variance Student’s t-test.

Clinical and demographic details of all patients and healthy controls are presented in Table 1 and include age, sex, body mass index (BMI), requirement for ventilation during admission and ICU admission levels of serum CRP, blood D-dimer levels and white blood cell counts along with their differentials. The average age of the ICU patients was 63.5 years with a 2-to-1 bias towards male patients, consistent with previous patient demographic reports linking more severe COVID-19 with older male patients (45). Table 1 also bins patients into two clinical outcome groups of “Short-Stay” and “Long-Stay/Died” based on the length of time in the ICU and survival: “Short-Stay” patients are classified as those spending < 6 days in the ICU and were survivors, while “Long-Stay/Died” patients are defined as patients who spent 6 or more days in the ICU or died during their stay in ICU. Combining patients who spent 6 or more days in the ICU with patients who died during their stay in ICU into a single clinical outcome category of “Long-Stay/Died” is based on the not unreasonable assumption that both sub-groups of patients can be defined here as having more severe COVID-19 than those spending < 6 days in the ICU and were survivors. The application and use of these two clinical outcome categories were more powerful in identifying immune differences between categories that the use of more simple categories of “survived versus died” or “did or did not require subsequent ventilation” (data not shown). The choice of 6 days as the cut-off was based upon iterative empirical statistical analyses of immune data: the cohorts were sequentially divided into two test sub-groups (for example, “<1 day” and “1+ days”, “<2 days” and “2+ days”, “<3 days” and “3+ days” etc.) and the immune data of the test subgroups were compared by Student’s t-test to identify the sub-groups with distinct clinical outcomes that had the greatest statistical significance (by p-value) with respect to differences in immune markers (Fig. 1A, C). While some patients were transferred to the ICU from a general ward rather than being directly admitted, there was no significant difference between the Short-Stay and Long-Stay/Died clinical outcome groups with respect to the mean days spent in a general ward prior to admission to ICU (p = 0.06).

**Table 1.** Cohort clinical information.

| Characteristics | Healthy (H)<br>(n = 8) | Cytokine Cohort<br>(n=90)* | Short-Stay (SS)<br>(n = 34) | Long-Stay/Died (LS/D)<br>(n = 56) |
| --- | --- | --- | --- | --- |
| Age (median years, min:max) | 53, 28:67 | 64, 28:95 | 56, 31:95 | 67, 28:92 |
| Sex (M:F) | 5:3 | 56:34 | 16:18 | 40:16 |
| Diagnosis | Asymptomatic/ healthy | SARS-COV-2 + | SARS-COV-2 + | SARS-COV-2 + |
| Collection timepoint (days) † | NA | 0-2 post admission | 0-2 post admission | 0-2 post admission |
| Severity | NA | ICU | ICU | ICU |
| BMI (median) | NA | 31 | 28 | 32 |
| CRP (median mg/L) | NA | 73.5 | 70 | 77 |
| D-dimer (median ng/ml) | NA | 1056 | 960.5 | 1248 |
| Length of ICU stay | NA | NA | <6 | ≥ 6 |
| Days in ICU (median, min:max) | NA | 7, 0:35 | 3, 0:5 | 11, 1:35 |
| Outcome (n; recovered:died) | NA | 74:16 | 34:0 | 40:16 |
| Mechanically ventilated (n, %) | NA | 45, 60 | 12, 38 | 33, 78 |
| PBMCs (median 10<sup>9</sup>/L) | 2.6 | 1.6 | 1.6 | 1.6 |
| PMNs (median 10<sup>9</sup>/L) | 3.6 | 7.7 | 7.1 | 8.2 |

\*BMI, D-dimer, CRP and ventilation information was not available for 16/90 patients; calculations were adjusted accordingly.
†71/90 samples were drawn at day 0, 17/90 at day 1 and 2/90 at day 2.
BMI: Body mass index; CRP: C-reactive protein; PBMCs: Peripheral blood mononuclear cells; PMNs: Polymorphonuclear leukocytes

All patients were equally treated with 6mg/day of Dexamethasone. Importantly, we found no significant differences between the two clinical outcome groups with respect to mean age, BMI, blood clotting parameters (D-dimer levels) or serum CRP levels (Fig. 1D, E). At admission, the mean total PMN counts were significantly increased in the Long-Stay/Died group compared to the healthy controls (p < 0.0001) and compared to the Short-Stay group (p = 0.025) (Fig. 1F). In our separate analyses of just the Initial Cohort and Replication Cohorts, however, differences in PMN counts were not statistically significant and thus we did not consider this measurement as a useful prognostic biomarker of clinical outcomes in the context of smaller cohort numbers. PBMC counts were also not significantly changed between healthy controls and patients or between our two clinica outcome groups (Fig. 1F). Thus, while these routine clinical tests follow a broad spectrum of parameters including inflammation, coagulopathy, hypo-immunity and autoimmunity, none consistently proves prognostic in identifying patients who would require an extended stay in the ICU or die. Accordingly, we conducted more detailed immunological examinations focused on a single, specific COVID-19-associated process, namely inflammation.

### Serum cytokine analyses as prognostic screens for predicting clinical outcome

We began by examining serum levels of IL-1β, IL-6, IL-10 and TNFα at ICU admission in all serum samples from our Cytokine Cohort of 90 COVID-19 patients. Strikingly, we found that the mean ICU admission levels of serum IL-10 (p = 0.004) and TNFα (p = 0.0005) were significantly elevated in the Long-Stay/Died group relative to the Short-Stay group (Fig. 1G). In the Cytokine Cohort, 43% (39/90) of patients had serum ICU admission levels of IL-10 levels ≥ 15pg/ml and 79% (31/39) of these patients fell into the Long-Stay/Died group. Similarly, 42% (38/90) of patients in the Cytokine Cohort had serum ICU admission levels of TNFα ≥ 10pg/ml and 79% (30/38) of these patients were members of the Long-Stay/Died group. Interestingly, serum IL-10 and TNFα only showed a weak correlation with each other (Pearson correlation coefficient R^2^ = 0.12, p = 0.27) suggesting that each may represent a different aspect or chronology of the inflammatory process. In parallel analyses of serum samples from 10 healthy controls (not matched for age or sex) the mean IL-10 level was 1.0 pg/ml (range 0.56 to 1.8) and the mean TNFα level was 2.4 pg/ml (range 1.7 to 3.9) (data not shown). Since the Cytokine Cohort represented a significant number of COVID patients (n = 90), a parallel replication cohort was not constructed.

Given the significant differences in ICU ***admission levels*** of TNFα (p = 0.0005) and IL-10 (p = 0.004) between the Short-Stay the Long-Stay/Died clinical outcome groups, we also examined the subsequent mean maximum serum cytokine levels in ***post admission*** samples from patients in the two groups and found an even more significant difference between the two groups for both serum TNFα (p < 0.0001) and serum IL-10 (p = 0.0009) (Fig. 1H). Intriguingly, many patients in the Long-Stay/Died group who demonstrated modest admission levels of serum IL-10 and TNFα subsequently developed high levels during their stay in ICU, further reinforcing the importance of these two cytokines as predictive measures of patient outcomes and monitoring the trajectory of disease.

While admission levels of serum IL-6 were also significantly different between the two clinical outcome groups (p = 0.007) (Fig. 1I), we excluded this cytokine from further analyses due to the potential confounding effects of anti-IL-6 receptor antibody (tocilizumab) treatments, which has routinely been administered to COVID-19 patients in British Columbia during ICU admission since February 2021 and such treatments could complicate the interpretation of our results. In this study, only six patients in the combined Initial and Replication cohorts received tocilizumab at the time of ICU admission (two patients in the Short-Stay group and four patients from the Long-Stay/Died group). These patients were not significant outliers with respect to the biomarkers of interest presented in this study and thus were not excluded from further analyses. Finally, there were no significant differences between the two clinical outcome groups with respect to mean serum IL-1β levels at ICU admission (p = 0.205) and thus this cytokine was also not analyzed further (Fig. 1I). In summary, we found that ICU admission levels of serum IL-10 and TNFα were useful and statistically powerful prognostic markers for clinical outcomes in severe COVID-19.

### Major PBMC subsets fail to distinguish Short-Stay from Long-Stay/Died patients

We examined whether parallel CyTOF analyses of peripheral immune cells sampled at the time of ICU admission could reveal additional prognostic biomarkers that identify patients in the Long-Stay/Died group, particularly among those that had serum IL-10 levels <15pg/ml and/or serum TNFα levels <10pg/ml. PBMC samples were available for 42/90 of the Cytokine Cohort patients and these 42 samples were divided into an Initial Cohort (14 samples) and a Replication Cohort (28 samples).

Using a 35-marker CyTOF panel on the Initial Cohort (Supplemental Figure 1) and a 38-marker CyTOF panel on the Replication Cohort (Fig 2.), we saw no differences between the Short-Stay patients and Long-Stay/Died patients with respect to major peripheral blood immune populations. The more focused and larger 38-marker CyTOF panel, used to analyze immune cell subsets in the Replication Cohort, permitted clear identification of broad blood cell lineages (B, T, NK and myelomonocytic) as well as major subsets within each cell lineage leading to 41 distinct clusters based on the variable expression of these cell-surface markers (Fig. 2A-C). A summary of mean absolute counts and p-values can be found in Supplementary Table 2. While these analyses of the Replication Cohort samples and the Initial Cohort samples confirmed previous reports (5, 46) of general lymphopenia in COVID-19 patients relative to healthy controls with respect to both total CD4 T cells and total CD8 T cells, these markers failed to discriminate between the Short-Stay and Long-Stay/Died patient groups. Also consistent with previous reports, we observed no significant differences in total B cells in COVID-19 patients compared to healthy controls or between the two clinical outcome groups (Fig. 2D). Although mean total NK cells, MAIT cells, γδ T cells, DC2/3 and pDC were depleted in COVID-19 patients relative to healthy controls these, too, failed to distinguish the Short-Stay group from the Long-Stay/Died group (Fig. 2E). Finally, while mean total monocytes and stem cell levels were significantly increased in COVID-19 patients relative to healthy controls, neither total monocyte levels nor total stem levels was able, individually, to distinguish the Short-Stay from the Long-Stay/Died patient groups (Fig. 2F, Table S2). In summary, broad immune subset analyses were insufficient to predict COVID-19 patient clinical outcomes with respect to the length of stay in ICU and/or death in either the Initial Cohort or the Replication Cohort. We, therefore, performed more detailed analyses of immune cell subsets within these broad cell categories to identify more subtle potential differences between the two clinical outcome groups that could assist in the prospective identification of Long-Stay/Died patients.

**Fig. 2.**
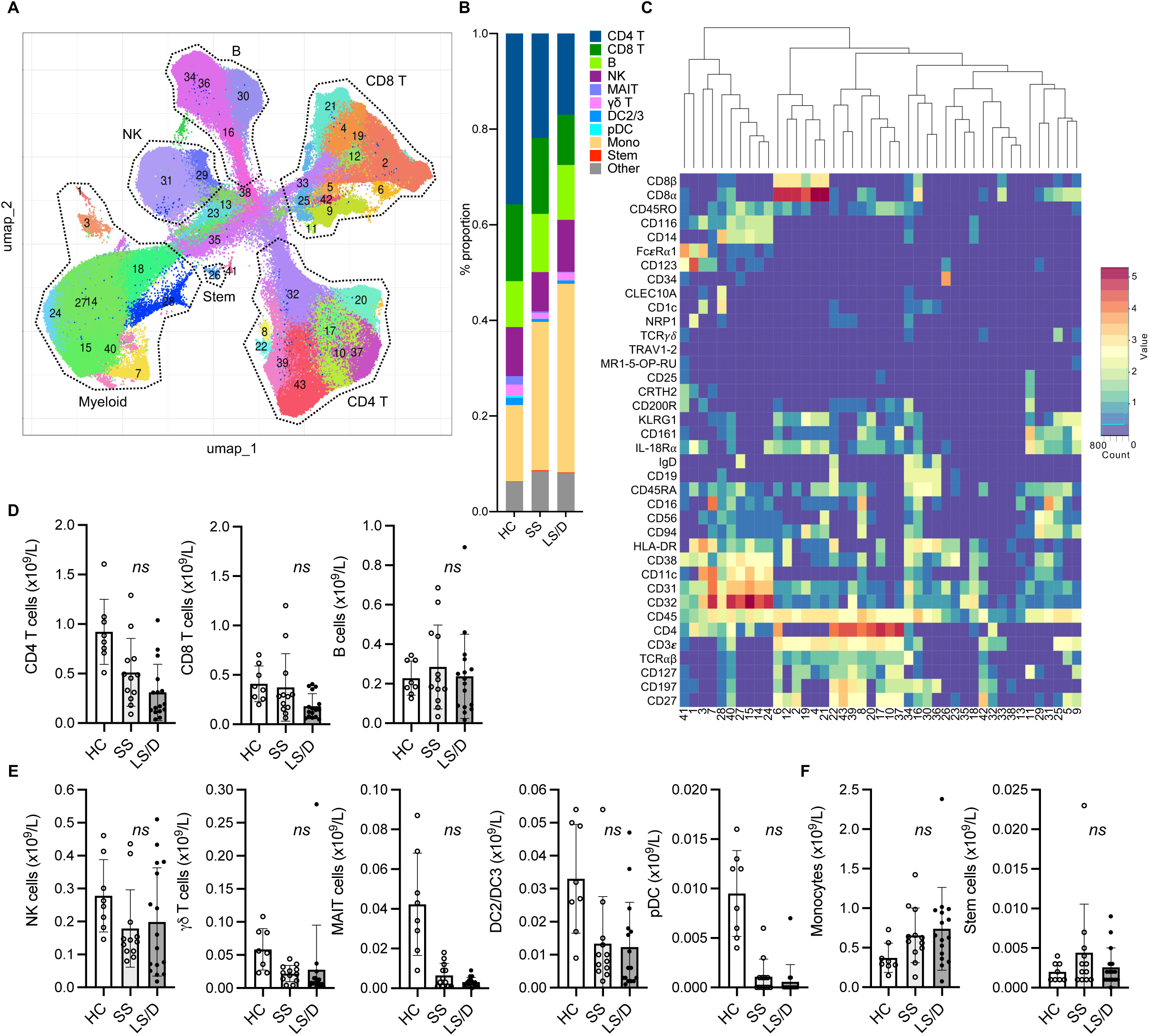
Major PBMC subsets fail to identify Long-Stay/Died patients (Replication Cohort). UMAP projection of ungated CyTOF-derived data from the replication cohort (n=28) (**A**). Proportion of immune cell subsets in Healthy Controls (HC), Short-Stay (SS) and Long-Stay/Died (LS/D) patient outcome groups (**B**). Mean marker expression heatmap of clusters shown in A (**C**). Absolute counts of adaptive PBMC subsets (CD4 T, CD8 T, B) (**D**), innate and unconventional subsets (NK, MAIT, γδ T, pDC, DC2/3) (**E**), monocytes and stem cells (**F**). *, p < 0.05; **, p < 0.01; ***, p < 0.001; ns, p ≥ 0.05 by two-tailed, two-sample unequal variance Student’s t-test.

### Levels of a distinct monocyte subset at the time of ICU admission predicts subsequent clinical outcome

To reveal a larger diversity of specific immune cell subsets we performed more focused cluster analyses on patient PBMC samples from the Initial Cohort and the Replication Cohort after pre-gating for selected major cellular subsets. To restrict the clustering to the myelomonocytic compartment we performed gated analyses on GM-CSFR^+^(CD116^+^) CD19^-^ CD3^-^ cells (Fig. 3A) and restricted clustering to shared marker channels to enable direct comparison between the Initial and the Replication cohorts. A summary of mean absolute cell counts and p-values can be found in Supplementary Table 3. These analyses did not reveal subsets that separated Short-Stay from Long-Stay/Died patients with respect to absolute cell counts. To focus more specifically on the monocytic subsets as well as to simplify the cluster analyses, after pre-gating on CD116+ CD19- CD3- cells, we restricted the marker channels selected for clustering to a set of 7 markers useful in defining monocytic subsets (CD45, CD14, CD16, CD11c, HLA-DR, CD123, CD56, see Figs. 3B, C). Interestingly, this strategy revealed a CD11c^low^ classical monocytic subset (CD45^+^ CD116^+^ CD3ε^-^ CD11c^low^ HLA-DR^+^ CD14^+^ CD16^-/low^ CD123^-/low^) that, in both the Initial and the Replication Cohorts, was consistently enriched in COVID-19 patients relative to healthy controls (Bonferroni-adjusted p = 0.006, Replication Cohort) and was preferentially enriched in the Long-Stay/Died group relative to the Short-Stay group (Bonferroni-adjusted p = 0.076, Replication Cohort) (Fig. 3D), though the latter did not achieve statistical significance. The potential prognostic value of the CD11c^low^ classical monocytic marker was restricted to this subset of classical monocytes in both the Initial and Replication Cohorts and did not reflect underlying changes of total classical monocytes which were unchanged in the two clinical outcome groups (Fig. 3E). Moreover, for both Initial and Replication Cohorts, total intermediate monocytes (CD14^+^ CD16^int^) and total non-classical monocytes (CD14^low^ CD16^+^), as well as observed subpopulations of these types of monocytes, did not prove useful prognostically (Fig. 3F). Focusing the analyses further on classical monocytes, we found that a three-marker gating strategy was sufficient to identify the CD11c^low^ classical monocyte population identified by our multi-dimensional analyses (shown here for one representative sample from each group in the Replication Cohort, where intensity is proportional to relative frequency of cells) (Fig. 3G). Thus, the prognostically useful biomarker of CD11c^low^ classical monocytes was detectable in two dimensions in both the Initial and the Replication Cohorts using antibodies to a small set of cell-surface markers.

**Fig. 3.**
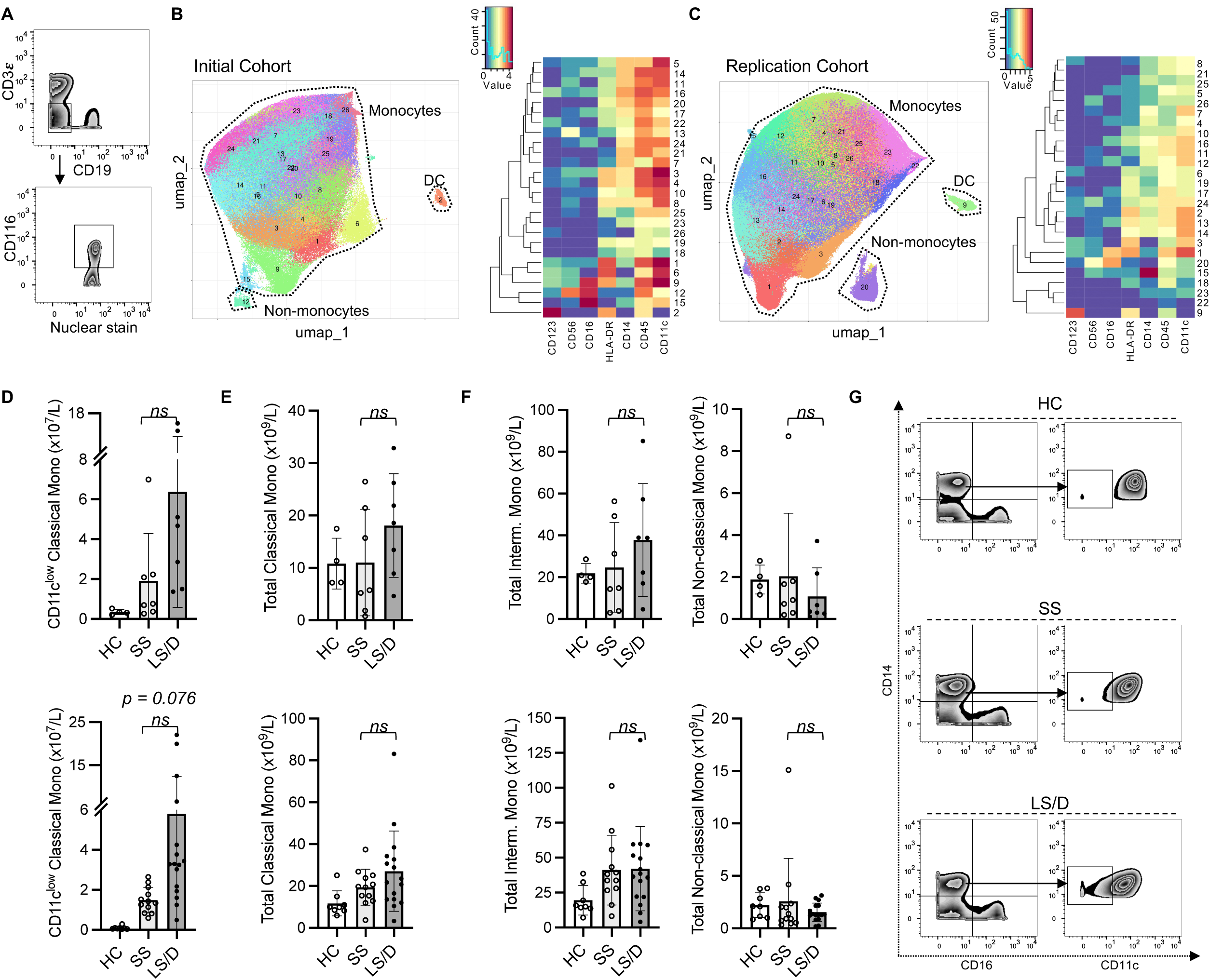
CD11c^low^ Classical Monocytes are predictive of clinical outcome. Representative gating of CD116^+^CD3^-^CD19^-^ cells (**A**). Initial Cohort UMAP projections of CD116+ CD3^-^ CD19^-^ gated cells (all samples combined; limited clustering channels) and mean marker expression heatmap of UMAP projections (**B**). Same as in B but for the Replication Cohort (**C**). Initial (top) and Replication Cohorts (bottom) absolute counts of monocyte subset predictive of clinical outcome (**D**). Absolute counts of non-significant monocyte subset identified based on gated clustering (top row: Initial Cohort, bottom row: Replication Cohort) (**E-F**). Manual gating strategy to view predictive monocyte subset (**G**). ns, p ≥ 0.05 by two-tailed, two-sample unequal variance Student’s t-test (with Bonferroni adjustment for multiple comparisons in (D)).

Because lymphopenia has been a consistent feature of severe COVID-19 and T cell subset alterations have been described, we performed similar in-depth analyses of the T cell compartments by gating on CD3^+^ cells prior to clustering. Consistent with our analyses of major subsets in the previous section, we were able to confirm and extend our and other groups’ findings that more subtle T cell subsets are significantly depleted in patients relative to healthy controls including subsets in the CD4^+^, CD8^+^, MAIT and γδ T cell compartments (Supplementary Figure 2). However, while we gained valuable insight into the altered T cell responses in COVID-19 patients, none of these T cell subsets was prognostically useful in separating the Long-Stay/Died patients from the Short-Stay patients.

### Combined evaluation of immune parameters as a tool to predict clinical outcome

Since deeper analyses of multiple cytokines and cell subsets at ICU admission revealed significant differences between the Long-Stay/Died and Short-Stay groups, we sought to combine these findings to generate a streamlined prognostic tool that could accurately predict whether a patient, newly admitted to ICU, was likely to have a subsequent longer stay in ICU or die. Although both serum TNFα and serum IL-10 were significantly elevated in Long-Stay/Died patients relative to Short-Stay patients in the Cytokine Cohort, using Pearson analyses, the length of stay in ICU correlated more significantly with serum IL-10 levels (R^2^ = 0.48, p < 0.001) and maximum IL-10 levels (R^2^ = 0.54, p < 0.001) than with serum levels of TNFα (R^2^ = 0.14, p = 0.19) (Supplementary Figure 3). Thus, we proceeded with only serum IL-10 as our cytokine-based pre-screen portion of a stepwise integrated prognostic tool. As the first step, using a cut-off value of 15pg/ml for serum IL-10 (data from the 90-sample Cytokine Cohort), demonstrated a 79% specificity and 55% sensitivity (positive likelihood ratio 2.6) in predicting that a patient newly admitted to ICU would have a longer stay in ICU or die (Fig. 4G). This prognostic specificity of 79% is somewhat comparable with that seen in the smaller subsets of the Cytokine Cohort, namely the 14-sample Initial Cohort (86%) and the 28-sample Replication Cohort (100%). Similarly, the prognostic sensitivity of 55% in the Cytokine Cohort is somewhat comparable with that observed in the Initial Cohort (86%) and the Replication Cohort (56%) (Figs. 4A-C, G). The variations in estimates of prognostic sensitivity and specificity between cohorts (the Cytokine Cohort and its two subsets of the Initial Cohort and Replication Cohort), however, likely demonstrate variations that reflect the influences of random patient sampling and, very importantly, cohort size. These results validate serum IL-10 levels as a pre-screen to identify patients likely to die or to experience a long ICU stay.

**Fig. 4.**
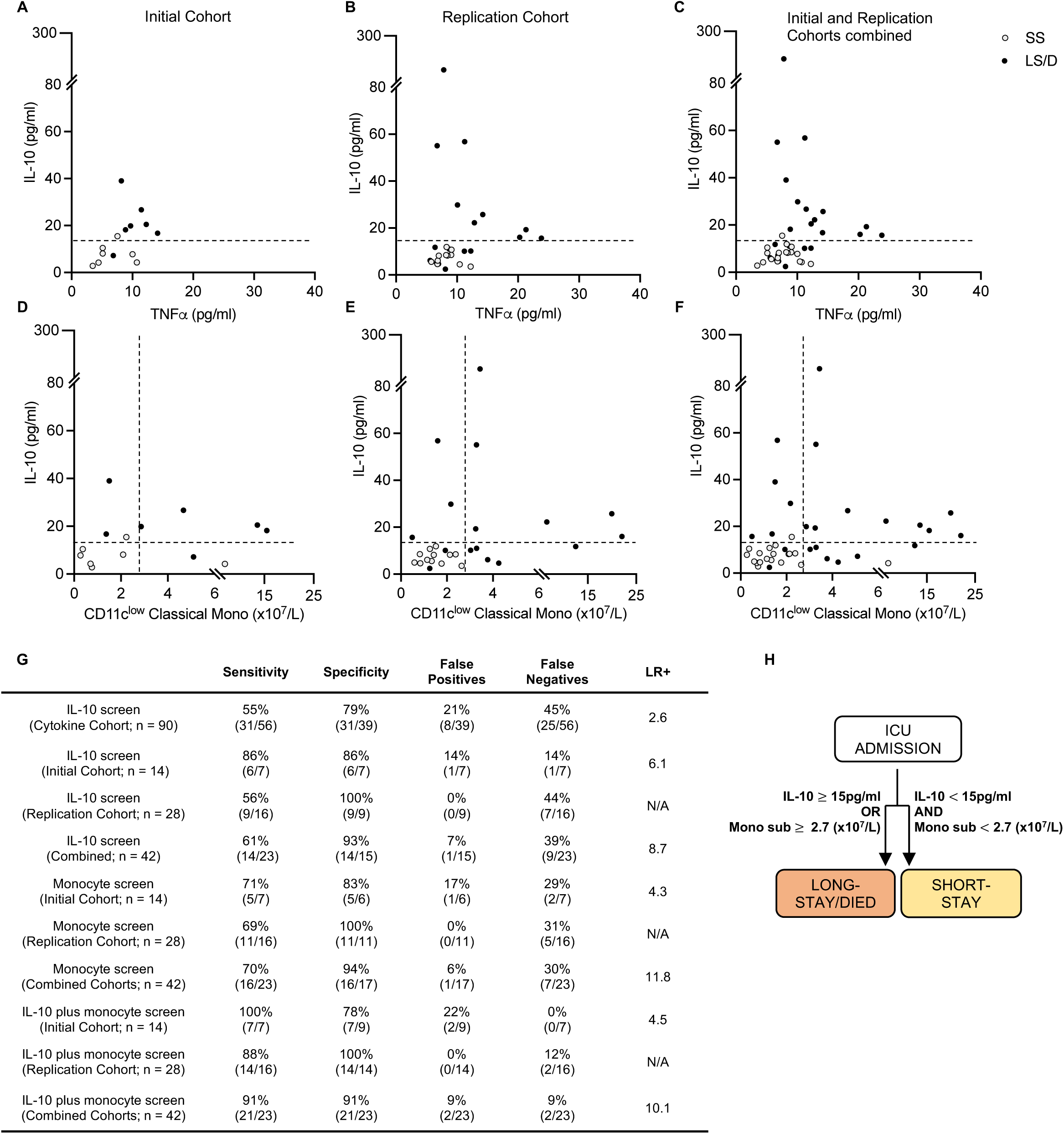
Prognostic cytokine and cellular biomarkers predict clinical outcome. Cytokine levels scatter plots of Initial (left), Replication (middle) and combined Cohorts (right) with dashed lines at cut-off value of 15pg/ml for serum IL-10 (**A-C**). Cytokine and cellular levels scatter plots for Initial (left) Replication (middle) and combined cohorts (right) with dashed lines at cut-off values of 15pg/ml and 2.7 (x10^7^/L) of serum IL-10 and CD11c^low^ classical monocytes respectively (**D-F**). Sensitivity, Specificity and Positive Likelihood Ratio (LR+) calculations for each screen and cohort (**G**). Prognostic patient screening chart based on serum IL-10 and CD11c^low^ monocyte subset measurement (**H**). *, p < 0.05; **, p < 0.01; ***, p < 0.001; ns, p ≥ 0.05 by two-tailed, two-sample unequal variance Student’s t-test and R^2^ by two-tailed Pearson correlation with 95% confidence interval. “Combined Cohorts” represent merged Initial and Replication Cohorts.

We then explored the utility of combining serum IL-10 levels (with a cut-off value of 15pg/ml) with levels of CD11c^low^ classical monocytes (with a cut-off value of 2.7×10^7^/L) as a stepwise integrated diagnostic tool. With this approach, 100% of the Long-Stay/Died patients were correctly identified in the Initial Cohort and 88% of Long-Stay/Died patients were correctly identified in the Replication Cohort, the latter with a specificity of 100% (Figs. 4D, E, G). These analyses of all 42 patients in the combined Initial and Replication Cohorts (n = 42) allowed us to predict with 91% sensitivity and 91% specificity (positive likelihood ratio 10.1) the clinical outcome of COVID-19 patients newly admitted to the ICU with respect to the likelihood of extended stay or death in the ICU (Figs. 4F, G). Thus, our results suggest that a simple screen of two biomarkers at the time of ICU admission allows for rapid identification of those patients who are likely to die or require extended ICU care (Fig. 4H) and has clear implications for patient care and health care delivery.

## Discussion

The goal of the present study was to identify prognostic biomarkers that, at time of ICU admission, could predict subsequent clinical outcome of COVID-19. Such markers are in urgent need and, with further testing and refinement, could serve to triage patients into specific groups for timely and appropriate care while, at the same time, offer insights into the immune-mediated determinants of disease response. Indeed, two patients (one in the Initial Cohort and one in the Replication cohorts) that were admitted to the ICU for less than one day may be examples of clinical mis-triage – using the prognostic biomarkers described here these patients would have been clearly identified as belonging to the < 6 Days clinical outcome group. Like many previous studies, we found that although severe COVID-19 is linked to broad shifts in peripheral blood immune subsets (increased PMNs and T cell lymphopenia) and increased blood inflammatory markers (CRP, D-dimer, etc.), none of these proved prognostic with respect to subsequent length of ICU stay and/or death. Therefore, we used CyTOF-based PBMC immunophenotyping and serum cytokine analyses on samples drawn at ICU admission to focus our attention on more subtle shifts in inflammatory parameters with a view to identifying prognostic biomarkers. Through iterative empirical analyses of these data, we identified two groups of ICU patients who would subsequently have clinically distinguishable disease outcomes: those who would be discharged from the ICU within 6 days and those who would require a longer ICU stay or would die. We then used retrospective analyses to generate a simple set of biomarkers that could easily be applied in the clinic to identify, at the time of ICU admission, those patients at greater risk of death or lengthy stay in the ICU. While the clinical decision to admit a patient to the ICU and indeed the length of stay considered clinically necessary for each patient may vary between ICUs, we sought to normalize these more subjective variables to the extent possible by limiting our patient cohort recruitment to only two ICUs, each ICU being located in a major teaching hospital of the same university.

A high admission level of serum IL-10 (≥ 15pg/ml), alone, was (in a cohort of 90 patients) a limited biomarker that identified patients in the Long Stay/Died group with 79% specificity, though with a lower sensitivity of 55% (positive likelihood ratio 2.6). Additional high dimensional cell surface protein analyses of 42 patients revealed a simple set of monocyte markers, specifically those identifying CD11c^low^ classical monocytes, that when combined with admission serum IL-10 levels accurately predicted with 91% specificity and 91% sensitivity (positive likelihood ratio 10.1) at the time of ICU admission patients who would subsequently either have a longer stay in ICU or who would die (validated in initial and replication cohorts). This prognostic power of combining two biomarkers into a single test was achieved despite the observation that the second biomarker (CD11c^low^ classical monocytes) alone was not statistically useful in distinguishing between the two clinical outcome cohorts (Bonferroni-adjusted p = 0.076) and provided only limited prognostic sensitivity (Fig. 4G). Thus, based on the information from our evaluation of 4 serum cytokines and 38 surface markers and validated on two separate clinical cohorts, we have distilled our prognostic screen down to a composite test of ***one cytokine and one monocyte subset*** as predictive biomarkers that could be evaluated in most clinical laboratories. Indeed, our demonstration that the cellular biomarker can be detected and visualized in two dimensional analyses (Fig. 3G) using limited markers reinforces the likelihood that this biomarker will be detectable using conventional clinical flow cytometry.

Although individually several of the biomarkers examined here have been investigated previously and described as markers of disease severity, there has been a lack of clear consensus on their prognostic utility in the published literature. For example, both IL-6 and IL-10 emerged early as candidate clinical markers of disease severity, but to our knowledge are not widely used in standard prognostic testing at hospital or ICU admission (32, 33, 36, 37). This likely reflects the fact that, used in isolation and without a detailed quantitative evaluation of threshold levels predicting outcome, their presence or absence provides a more superficial indication of current inflammatory status and fails to predict the temporal trajectory of clinical disease (increasing or decreasing severity). This also may explain why anti-IL-6 receptor therapy has shown only limited efficacy as a broad-spectrum therapeutic for severe COVID-19 and fails to reduce overall mortality (13, 47). Similarly, although corticosteroids have emerged as a standard-of-care for COVID-19 ICU patients and undoubtedly provide improved recovery after infection, they are widely recognized as “double-edged swords”: while they are effective at suppressing excessive inflammation, they also potently suppress adaptive immune responses, potentially reducing viral clearance and increasing susceptibility to secondary infections (12, 48). With that backdrop, a benefit of the streamlined ICU biomarker panel described here is that it provides a direct prognostic link to patient outcome and may also serve as a biomarker panel for monitoring patient response to therapies, though longitudinal analyses of these markers have not been provided here to measure either changes with resolving disease severity or response to therapies. Interestingly, a genetic association of variants of the *IL10Rb* gene with critical COVID-19 was recently identified through whole genome sequencing (49). This finding, together with our finding of the association of high levels of serum IL-10 with death and/or long ICU stay, may point to the important involvement of the IL-10 ligand/receptor axis in the evolution of severe COVID-19.

In our study, deep immunophenotyping of the myeloid compartment in COVID-19 patients proved pivotal in defining markers to predict patient outcomes. While we saw no significant early changes in total monocyte numbers or total classical monocyte numbers or frequencies, a prognostically useful monocytic subset was contained within these broader subsets which highlights the need for a high-dimensional evaluation to identify subtle, but informative, changes in immune subpopulations that might otherwise have been overlooked. After identification of such subtle biomarkers using high-dimensional analytic technologies, simpler two-dimensional technologies (using limited markers) can then be used to measure the biomarker clinically. It is noteworthy that previous studies have linked both overall increased numbers of inflammatory macrophages in the lung and increases of specific subpopulations of peripheral blood monocytes to severity of COVID-19 (50–53). In fact, Schulte-Schlepping *et. al.* specifically showed the accumulation of CD11c^low^ monocytes in severe COVID-19 patients (17). Other studies also reported subtle, monocyte subset-specific changes in severe COVID-19, including dysfunctional pro-inflammatory cytokine production, reduction in HLA-DR transcripts, accumulation of HLA-DR^low^ monocytes and reduction of non-classical monocytes (17, 34, 38, 54). The data presented here confirm and extend these observations in a manner that facilitates accurate prognostication. They also reveal CD11c^low^ classical monocytes as new target populations for more focused mechanistic studies in future research. While the combination of these two biomarkers certainly provides prognostic information on disease outcome in COVID-19, there is a possible parallel interpretation of the results: since all patients received corticosteroids at the time of admission, the biomarkers described here may actually be identifying those patients who are, in fact, more responsive to corticosteroid therapy. We leave this intriguing possibility for future investigation.

Although not specifically addressed here, we believe that these prognostic biomarkers provide a roadmap for future studies aimed at guiding and monitoring response to therapy. Such monitoring is particularly important in the context of therapies that have the acknowledged potential of exacerbating clinical disease if given in a temporally inappropriate manner in the COVID-19 cycle of stimulation and progression to clearance and resolution. While we have focused here on the utility of these markers at the time of ICU admission it is possible that these may prove even more valuable as temporal monitoring tools for revealing disease trajectory on this continuum and responses to therapeutic intervention.

## Materials and Methods

### Study design

This study was approved by the University of British Columbia Clinical Research Ethics board (H20-00685) and patient blood was collected at St. Paul’s Hospital and Vancouver General Hospital in Vancouver, BC following informed consent. All COVID-19 patients had a positive nasal or tracheal real time reverse transcription polymerase chain reaction (RT-PCR) SARS-CoV-2 test. To avoid unnecessary virus exposure, patient blood was collected in combination with routine care. Patient samples (n = 90) were collected within 48 hours of ICU admission between November 2020 and February 2021. Patient demographics and clinical information are listed in Table 1. Patient samples were transported to the main campus of the University of British Columbia for further processing. Healthy control blood samples (n = 8) were collected from age-matched volunteers who showed no COVID-19 symptoms or other illnesses and who had no history of COVID-19.

### Specimen collection and isolation

Blood designated for peripheral blood mononuclear cell (PBMC) analysis was collected into citrate-coated BD Vacutainer™ Glass Mononuclear Cell Preparation (CPT) tubes and PBMCs were isolated within four hours following collection according to manufacturer guidelines. Red blood cell lysis was performed with ACK lysing buffer (Gibco) for 10 minutes. Isolated PBMCs were frozen in fetal bovine serum (Gibco) with 10% dimethyl sulfoxide and stored in liquid nitrogen. Blood designated for serum analysis was collected into BD Vacutainer™ Serum Separation Tubes (SST) and allowed to clot for at least 30 minutes prior to centrifugation at 1200 rcf for 10 minutes and serum collection and storage at −80 °C.

### Antibody staining and CyTOF data collection

Frozen PBMCs were thawed at 37 °C and washed with RPMI 1640 containing 10% FBS and 25U nuclease (Thermo Fisher Scientific, Cat. #88700). Between 1-4 x 10^6^ cells per sample were used for antibody staining. Prior to fixation, all centrifugation steps were performed at 500 rcf and 4 °C. All reagent dilutions were prepared according to manufacturer instructions unless stated otherwise. For live/dead cell analysis, PBMCs were stained with Cell-ID™ Intercalator-Rh (Fluidigm, Cat. #201103A) for 15 minutes at 37 °C and washed with MaxPar® MCSB. Prior to surface staining, cells were incubated with human TruStain FcX™ (Biolegend, Cat. #422302) for 15 minutes at 4 °C and stained with a surface antibody cocktail for 30 minutes at RT (see Supplemental Table 2 for complete list of antibodies). The MR1-5-OP-RU tetramer was incubated together with the antibody cocktail. After incubation, the cells were washed and incubated for 30 minutes at RT with the secondary anti-APC antibody. Prior to fixation and nuclear staining, PBMCs were washed with MaxPar® MCSB (Fluidigm, Cat. #201068) and incubated in MaxPar® Fix and Perm Buffer (Fluidigm Cat. #201067) and Cell-ID™ Intercalator-IR (Fluidigm, Cat. #201192A) for 1 hour. Post fix, all centrifugation steps were performed at 900 rcf and 4 °C. To prepare for CyTOF acquisition, PBMCs were washed with MilliQ water and resuspended in EQ™ Four Element Calibration Beads (Fluidigm, Cat. #201078). Samples were acquired with a CyTOF®2 mass cytometer. An average of 400,000 events were collected for each sample at a flow rate of 45µl/min.

### Cytokine data collection

Serum cytokines IL-6, IL-10, TNFα and IL-1β were quantified using the Simoa HD-1 platform from Quanterix (Billerica, MA) according to manufacturing guidelines and as specified by Stukas *et. al.* (55).

### Data processing

All data files were normalized (https://github.com/nolanlab/bead-normalization) and events of interest were manually gated with the FlowJo gating software (BD Biosciences). Dimensionality reduction and clustering were performed with Uniform Manifold Approximation and Projection (UMAP) and Rphenograph respectively, as provided in the bioconductor package Cytofkit (https://github.com/JinmiaoChenLab/cytofkit2). The input files were equally down sampled and cytofAsinh was used as the transformation method. The Rphenograph k was set to the default of 30. The dimensionality reduction and clustering were both performed on the entire data set as well as separately on manually pre-gated populations. The assignment of clusters to specific immune cell subsets based on surface marker expression was guided by the cell-type definitions described in Supplementary Table 4.

### Statistical analysis and figures

Sample size and statistical tests are indicated in figure legends and all graphs and statistical tests were generated using GraphPad Prism (GraphPad Software, La Jolla California, USA). A test was considered statistically significant at a probability of < 5% (p < 0.05) and we did not assume a Gaussian distribution. Where indicated, the p-values for independent t-tests were adjusted for multiple comparisons (Bonferroni). Error bars represent mean ± SD. UMAP plots and heatmaps were exported from Cytofkit and experimental outline figures, including the graphical abstract were created using BioRender.com. Figures were assembled in Microsoft PowerPoint.

## Data Availability

All data produced in the present study are available upon reasonable request to the authors.

## Acknowledgments

We thank the healthcare teams from St. Paul’s Hospital and Vancouver General Hospital as well as the technical support from the Biomedical Research Centre core facility members Michael Williams (AbLab) and Andy Johnson (ubcFLOW cytometry). We acknowledge that the following reagent was obtained through the NIH Tetramer Core Facility: APC coupled MR1-5-OP-RU tetramer. Finally, we thank Jeff Biernaskie and Bryan Yipp at the University of Calgary for helpful discussions and generous seed funding from the Thistledown Foundation.

## Data and materials availability

All data produced in the present study are available upon reasonable request to the authors.

## Supplementary Information for

**Fig. S1.**
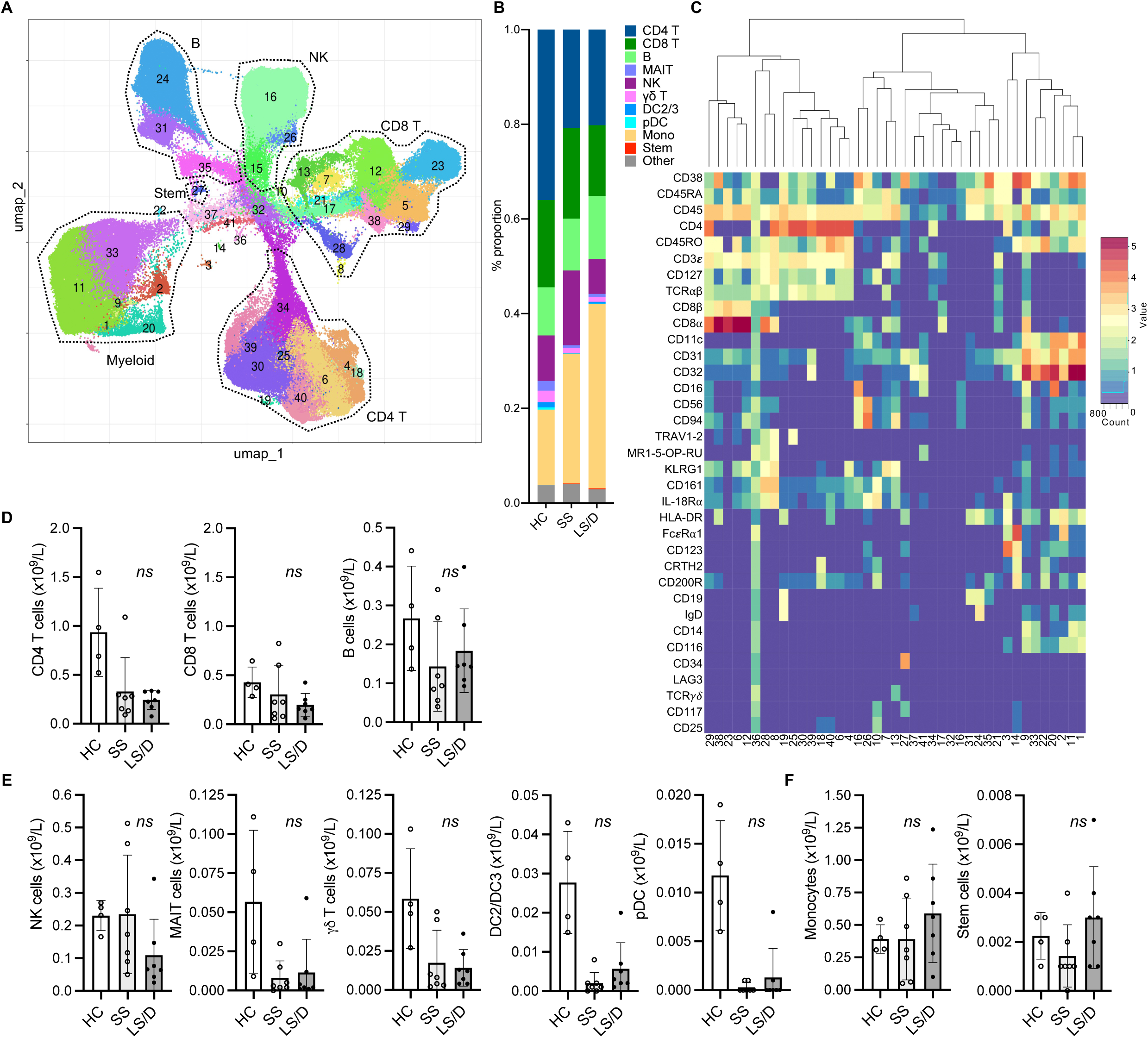
Major PBMC subsets fail to identify Long-Stay/Died patients (Initial Cohort). UMAP projection of ungated CyTOF-derived data from the Initial cohort (n=14) (**A**). Proportion of immune cell subsets in Healthy Controls (HC), Short-Stay (SS) and Long-Stay/Died (LS/D) patient outcome groups (**B**). Mean marker expression heatmap of clusters shown in a (**C**). Absolute counts of adaptive PBMC subsets (CD4 T, CD8 T, B) (**D**). Innate and unconventional subsets (NK, MAIT, γδ T, pDC, DC2/3) (**E**). Monocytes and stem cells (**F**). *, p < 0.05; **, p < 0.01; ***, p < 0.001; ns, p ≥ 0.05 by two-tailed, two-sample unequal variance Student’s t-test.

**Fig. S2.**
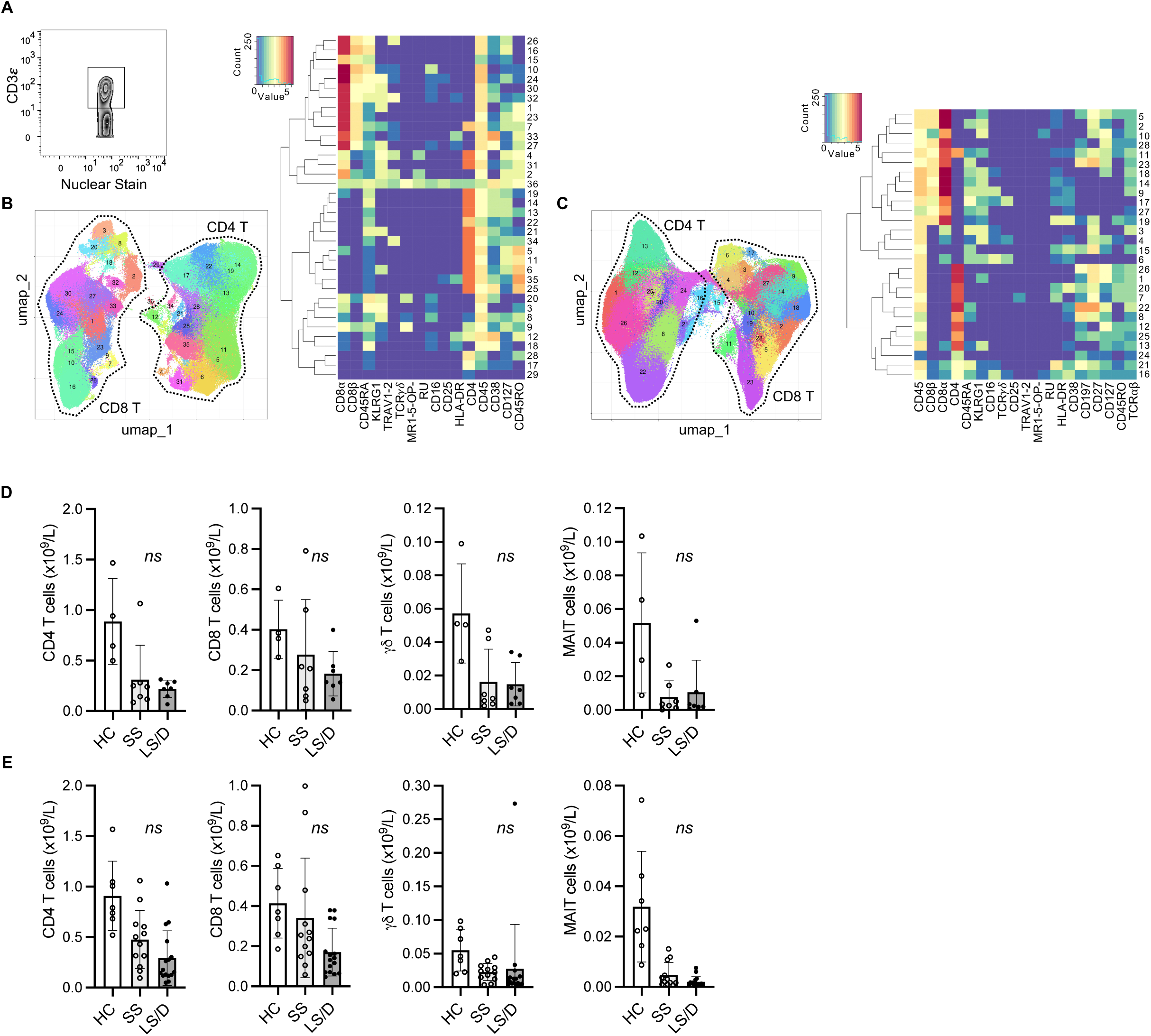
T cell subsets fail to identify Long-Stay/Died patients. Representative gating of CD3^+^ cells (**A**). Initial Cohort UMAP projections of CD3^+^ gated cells (all samples combined; limited clustering channels) and mean marker expression heatmap (**B**). Same as in B but for the Replication Cohort (**C**). Initial Cohort absolute counts of T cell subsets identified based on gated clustering (**D**). **S**ame as in D but for the Replication Cohort (**E**). *, p < 0.05; **, p < 0.01; ***, p < 0.001; ns, p ≥ 0.05 by two-tailed, two-sample unequal variance Student’s t-test.

**Fig. S3.**
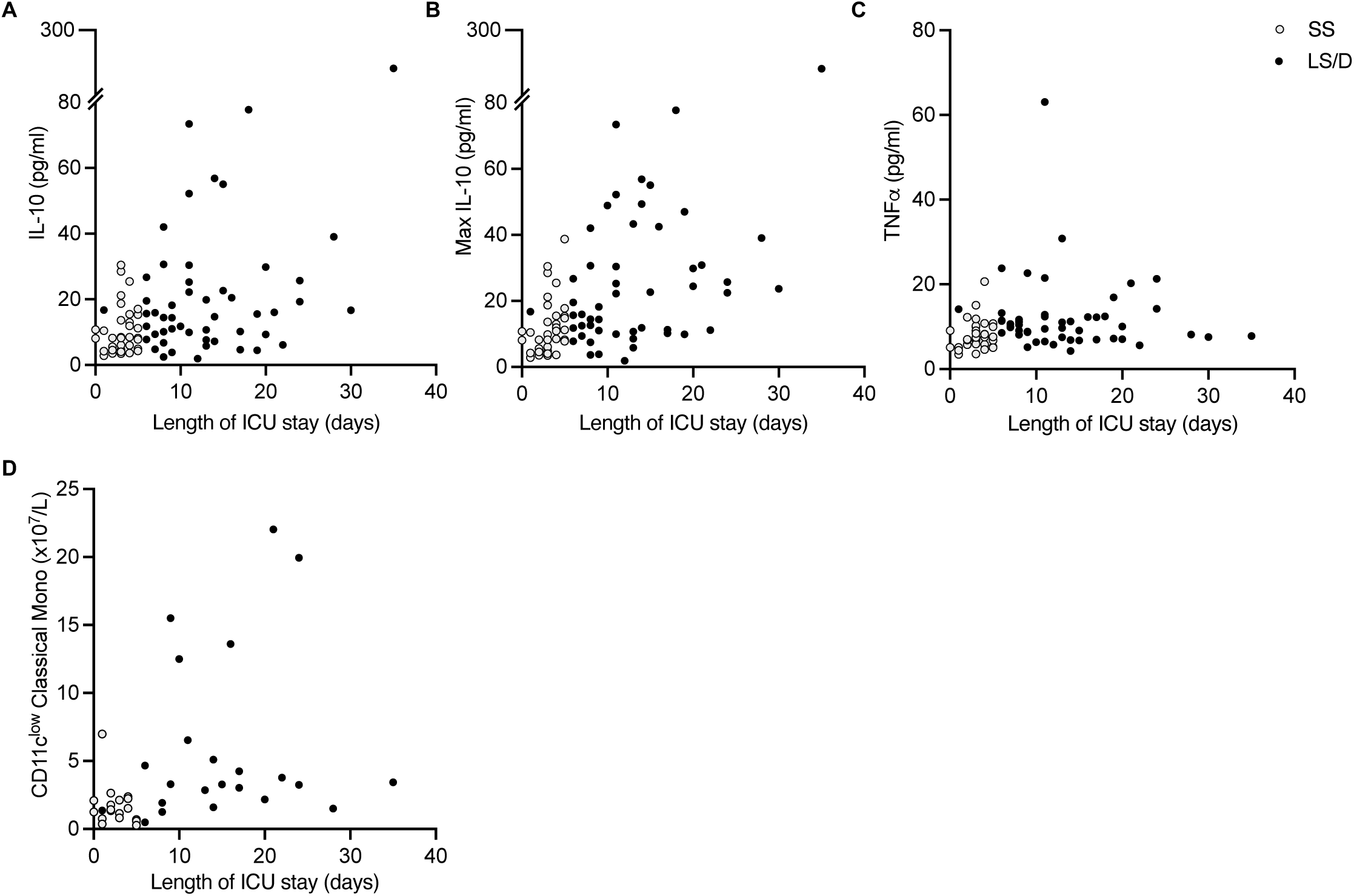
Cytokine and length of ICU stay correlations. Correlations of length of ICU stay with ICU admission serum IL-10 levels (**A**), maximum serum IL-10 during ICU stay (**B**) serum TNFα ICU admission levels (**C**) and CD11c^low^ Classical Monocytes (**D**). *, p < 0.05; **, p < 0.01; ***, p < 0.001; ns, p ≥ 0.05 by two-tailed, two-sample unequal variance Student’s t-test and R^2^ by two-tailed Pearson correlation with 95% confidence interval.

**Table S1.**
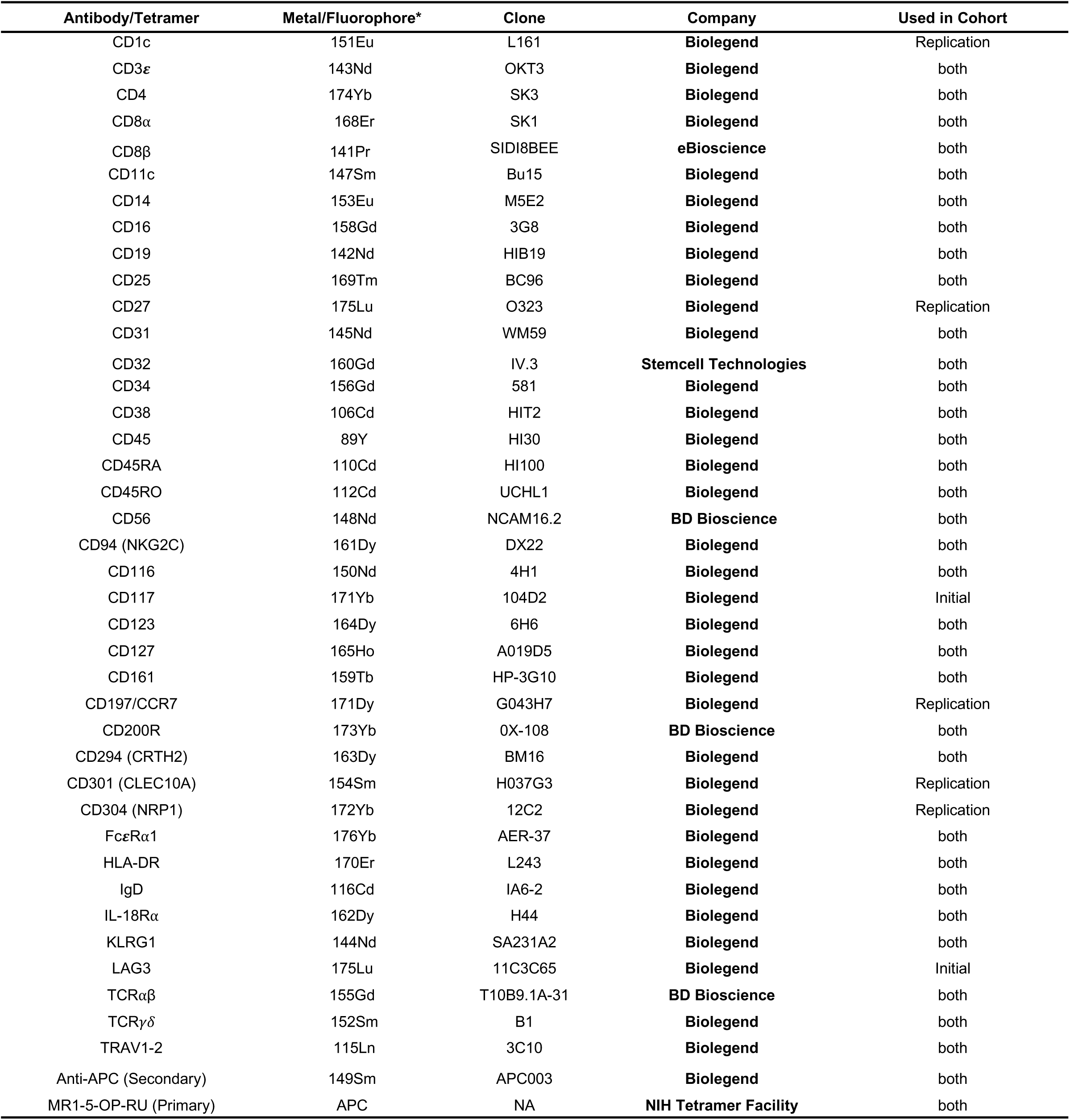
List of CyTOF antibodies.

**Table S2.** Mean absolute counts and p-values (ungated cluster analyses)

| Cohort | Total cell subset | H | SS | LS/D | H vs SS | H vs LS/D | SS vs LS/D |
| --- | --- | --- | --- | --- | --- | --- | --- |
| Mean absolute counts (x10<sup>9</sup>/L) |  |  |  |  | p-values |  |  |
| Initial | CD4 T cells | 0.936 | 0.330 | 0.244 | 0.067 | 0.053 | 0.549 |
|  | CD8 T cells | 0.428 | 0.304 | 0.198 | 0.383 | 0.049 | 0.402 |
|  | B cells | 0.267 | 0.143 | 0.184 | 0.176 | 0.335 | 0.509 |
|  | Monocytes | 0.392 | 0.392 | 0.589 | 0.997 | 0.237 | 0.311 |
|  | Stem cells | 0.002 | 0.001 | 0.003 | 0.247 | 0.315 | 0.088 |
|  | NK cells | 0.230 | 0.234 | 0.109 | 0.961 | 0.032 | 0.150 |
|  | MAIT cells | 0.057 | 0.008 | 0.012 | 0.122 | 0.140 | 0.721 |
|  | γδ T cells | 0.058 | 0.018 | 0.014 | 0.077 | 0.065 | 0.705 |
|  | pDC | 0.012 | 0.000 | 0.001 | 0.029 | 0.028 | 0.473 |
|  | DC2/3 | 0.028 | 0.002 | 0.006 | 0.026 | 0.034 | 0.205 |
| Replication | CD4 T cells | 0.911 | 0.511 | 0.309 | 0.008 | <0.001 | 0.112 |
|  | CD8 T cells | 0.386 | 0.374 | 0.184 | 0.914 | 0.005 | 0.090 |
|  | B cells | 0.247 | 0.285 | 0.237 | 0.584 | 0.871 | 0.565 |
|  | Monocytes | 0.403 | 0.657 | 0.740 | 0.041 | 0.028 | 0.618 |
|  | Stem cells | 0.002 | 0.004 | 0.003 | 0.223 | 0.451 | 0.360 |
|  | NK cells | 0.259 | 0.179 | 0.199 | 0.108 | 0.269 | 0.708 |
|  | MAIT cells | 0.047 | 0.007 | 0.003 | 0.001 | 0.000 | 0.076 |
|  | γδ T cells | 0.058 | 0.022 | 0.028 | 0.003 | 0.130 | 0.732 |
|  | pDC | 0.009 | 0.001 | 0.001 | <0.001 | <0.001 | 0.575 |
|  | DC2/3 | 0.037 | 0.014 | 0.012 | 0.003 | 0.001 | 0.829 |

p-values calculated using a two-tailed, two-sample unequal variance Student's t-test. H = Healthy; SS = Short-Stay; LS/D = Long-Stay/Died

**Table S3.** Mean absolute counts and p-values (gated monocyte and T cell cluster analyses)

| Cohort | Monocyte subset | H | SS | LS/D | H vs SS | H vs LS/D | SS vs LS/D |
| --- | --- | --- | --- | --- | --- | --- | --- |
| Mean absolute counts (x10<sup>9</sup>/L) |  |  |  |  | p-values |  |  |
| Initial | CD11c<sup>low</sup> Classical | 0.004 | 0.019 | 0.082 | 0.536* | 0.133* | 0.388* |
|  | Total Classical | 0.108 | 0.110 | 0.181 | 0.620 | 0.136 | 0.212 |
|  | Total Intermediate | 0.218 | 0.247 | 0.377 | 0.939 | 0.234 | 0.338 |
|  | Total Non-Classical | 0.019 | 0.020 | 0.011 | 0.844 | 0.080 | 0.462 |
|  | CD4 T cells | 0.888 | 0.312 | 0.219 | 0.010 | 0.004 | 0.506 |
|  | CD8 T cells | 0.403 | 0.277 | 0.182 | 0.309 | 0.005 | 0.415 |
|  | MAIT cells | 0.052 | 0.008 | 0.011 | 0.014 | 0.018 | 0.724 |
|  | γδ T cells | 0.057 | 0.016 | 0.015 | 0.009 | 0.006 | 0.868 |
| Replication | CD11c<sup>low</sup> classical | 0.001 | 0.015 | 0.058 | <0.001* | 0.010* | 0.076* |
|  | Total Classical | 0.117 | 0.193 | 0.271 | 0.035 | 0.010 | 0.167 |
|  | Total Intermediate | 0.196 | 0.412 | 0.422 | 0.025 | 0.021 | 0.925 |
|  | Total Non-Classical | 0.022 | 0.026 | 0.015 | 0.890 | 0.061 | 0.404 |
|  | CD4 T cells | 0.899 | 0.477 | 0.292 | 0.004 | 0.061 | 0.098 |
|  | CD8 T cells | 0.412 | 0.341 | 0.171 | 0.489 | <0.001 | 0.083 |
|  | MAIT cells | 0.035 | 0.005 | 0.002 | 0.001 | <0.001 | 0.093 |
|  | γδ T cells | 0.055 | 0.022 | 0.028 | 0.003 | 0.140 | 0.769 |

p-values calculated using a two-tailed, two-sample unequal variance Student's t-test.
H = Healthy; SS = Short-Stay; LS/D = Long-Stay/Died
\*p-values that have been Bonferroni-adjusted for multiple comparisons

**Table S4.**
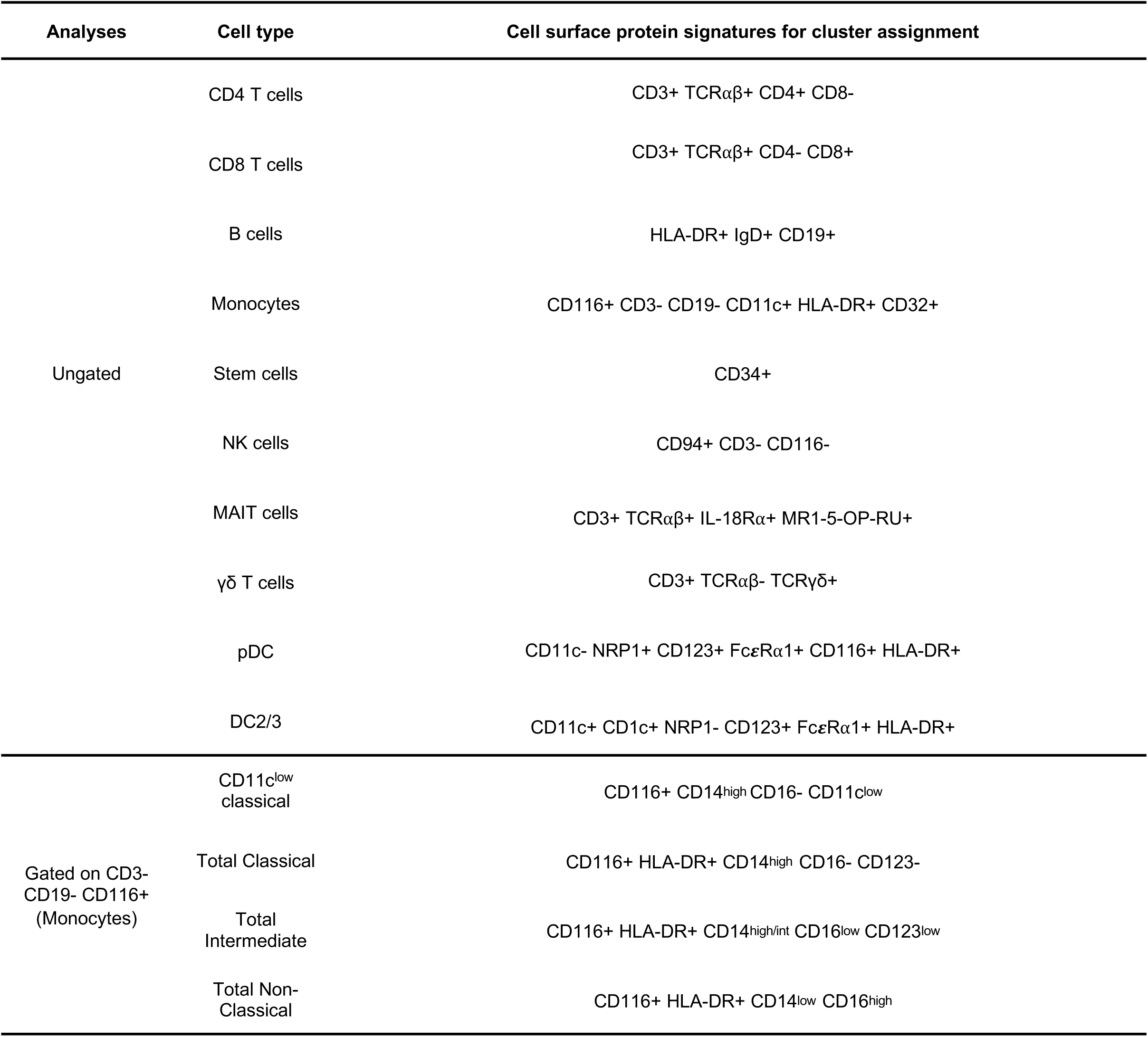
Cell surface protein signatures for cluster assignment.

